# Understanding the CoVID-19 pandemic Curve through statistical approach

**DOI:** 10.1101/2020.04.06.20055426

**Authors:** Ibrar ul Hassan Akhtar

## Abstract

Current research is an attempt to understand the CoVID-19 pandemic curve through statistical approach of probability density function with associated skewness and kurtosis measures, change point detection and polynomial fitting to estimate infected population along with 30 days projection. The pandemic curve has been explored for above average affected countries, six regions and global scale during 64 days of 22nd January to 24th March, 2020. The global cases infection as well as recovery rate curves remained in the ranged of 0 – 9.89 and 0 – 8.89%, respectively. The confirmed cases probability density curve is high positive skewed and leptokurtic with mean global infected daily population of 6620. The recovered cases showed bimodal positive skewed curve of leptokurtic type with daily recovery of 1708. The change point detection helped to understand the CoVID-19 curve in term of sudden change in term of mean or mean with variance. This pointed out disease curve is consist of three phases and last segment that varies in term of day lengths. The mean with variance based change detection is better in differentiating phases and associated segment length as compared to mean. Global infected population might rise in the range of 0.750 to 4.680 million by 24^th^ April 2020, depending upon the pandemic curve progress beyond 24th March, 2020. Expected most affected countries will be USA, Italy, China, Spain, Germany, France, Switzerland, Iran and UK with at least infected population of over 0.100 million. Infected population polynomial projection errors remained in the range of −78.8 to 49.0%.

## Introduction

Zoonotic coronaviruses are grouped into four genera of Alphacoronavirus, Betacoronavirus, Gammacoronavirus and Deltacoronavirus based on difference on polyprotiens, structural and accessory proteins^1^. The severe acute respiratory syndrome (SAR) coronavirus belongs to Betacoronavirus genus and has capability to spill over in term of its shift from natural host (bat, mouse) to intermediate host of other animals (camels, cats, cows, pigs) through genetic diversity and natural co-evolution^1,2^. The current pandemic of coronavirus (SARs-CoVID-2) referred as CoVID-19, not first time for the world, has threaten socio-economic deprivation and setback to UN sustainable development goals (SDGs). The H1NI influenza pandemic of 1918 provided us an insight related to deadliest history that had killed approximately 50–100 million population^3^. Some of answers to anticipatory questions are (a) virus origin (b) nature of pathogenesis (c) age relevancy with death (d) multiple year pandemic waves (e) do pandemic has predictable cycle? and possible future scenario to tackle such pandemics among developed as well as developing nations^3^.

There are certain aspects to be considered for any disease in relation to environment; to differentiate between actual association or causation based on its strength, consistency, specificity, temporality, biological gradient, plausibility, coherence, experiment and analogy^4^. The pandemic mitigation measures might include (a) an effective use of available data and experience (b) logical assessment of mitigation measures and (c) socio-economic and political assessments like isolation of sick peoples, self-isolations, change in daily life habits, travel restrictions, prohibition on social gatherings, educational institutions closures, use of masks and personal protective equipment’s, social distancing and vice versa^5^. An effective response towards pandemic demands an integrated approach of medical science with social science, data science, artificial intelligence, information technology, statistics, meteorology, biotechnology, biomedical science, anthropology, public health, diplomacy, logistics, crisis management and so on^6^.

No doubt, epidemic history offers considerable advice, if people knows history and respond with wisdom^7^. Data compiling, sharing and mapping provide real time approach to understand the temporal, spatial patterns and help decision and policy makers, health care specialist and social community like web based dashboard^8^. The Lancet (2020) highlighted that data openness, accuracy, reliability, sharing is of paramount for better response towards global health emergency like CoVID-19^9^. Infectious disease like CoVID-19 encourage a peculiar need to develop effective control plan based on epidemic trends and associated impacts on community^10^. The Casella (2020) explored the daily CoVID-19 pandemic data based draconian and nudge policies from control theory for China, Italy and UK^11^. The CoVID-19 pandemic critical characteristics includes fast unstable dynamics, measurement and actuation delay, uncertainty due to lack of knowledge and virus behaviour, lack of effective vaccines and poor health system response.

Time series data analysis of MERS-CoV based on trends, seasonality and statistical forecasting demonstrated use of underlying dynamics by healthcare policy makers and experts^12^. The Deb (2020) implemented autoregressive integrated moving average (ARIMA) method to explore CoVID-19 incidence pattern and disease reproduction number and provided understanding for current outbreak^13^. The disease analysis models like an improved SIR (suspected, infected and recovered) model of asymptomatic CoVID-19 provided an insight into northern Italy and outperformed standard SIR model^14-16^. A SIDHARTHE (Susceptible, Infected, Diagnosed, Healed, Ailing, Recognised, Threatened and Extinct) model also showed capability to discriminate patients among diagnosed and non-diagnosed cases for Italy. Identification of CoVID-19 cases is quickly possible through integration of mobile based survey and artificial intelligence that will reduce disease spread among high risk population^17^. While, the growth models based on logistics, Weibull and Hill equation delivered insight to statistical nature of CoVID-19 epidemiology^18^. Two other models of log-linear for percent change and linear for unit change are developed as data driven approach to understand the CoVID-19 and its gender associations in Hong Kong^19^. An interesting model that has been developed is to carry out global CoVID-19 risks assessment using four Chinese cities modelled fight data (passenger’s destinations) from FLIRT database^20^. Some studied targeted environmental parameters of temperature, relative humidity and wind speed to highlight underlying relationships and are statistically (Pearson correlations) linked with CoVID-19 spread and founds temperature to be negligible to moderately relevant^21^.

The outbreak cases isolation and expected contact tracing can only be feasible for reducing the size of pandemic and control over longer time period based on branching process model^22^. Current study is an attempt to understand the CoVID-19 pandemic curve based on statistical time series approach of daily disease rates, probability density function along with measures of its dispersion, change point detection and polynomial curve fitting and projection at global, regional and globally above average affected countries except for Pakistan.

## Method

The confirmed and recovered cases data are compiled from the daily situation reports downloaded from WHO organization for the period of 22nd January to 24th March, 2020^23^. These cumulative data is changed to daily data by subtracting of today value from the yesterday. All dataset is used to compute cases infection rate (CIR), cases recovery rate (CIR), Probability density functions (PDF), change point detection (CPD), 2nd to 4th order polynomial curve fitting along with 30 days projection (24th April, 2020) and evaluation of polynomial curve fitting based on percent change difference between actual and projected cases on 1st April, 2020.

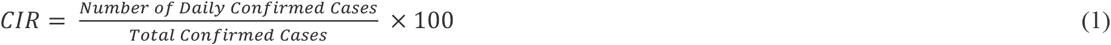

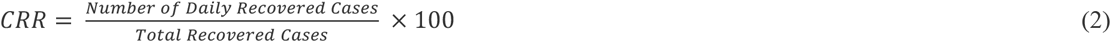

The kernel smoothing density generates PDF estimate along with measure of dispersion by skewness and kurtosis value for the confirmed and recovered cases in the variable vector^24-28^.

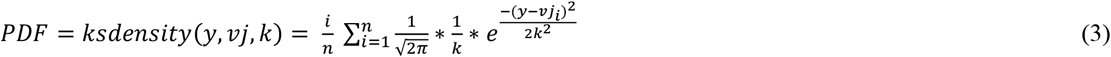

Where *y* is the value to be computed for density, *vJ* is the distributed sample used as kernel median, *k* (> 0) is the bandwidth used as scale of the kernel, and *PDF* is the required probability density estimate.

To understand the daily time series curve of CoVID-19 data, we used the multiple CPD based on binary segmentation method^29-31^ using Equation (4). Two methods are used for the CPD i.e., (i) change in mean using likelihood ratio and cumulative sum test statistics^32,33^ and (ii) changes in mean and variance using normal distribution^34^.

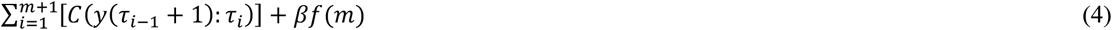

where *C* is a cost function for a segment e.g., negative log-likelihood and *βf*(*m*) is a penalty to guard against over fitting (a multiple change point version of the threshold c). A brute force approach to solve this minimization considers 2^n−1^ solutions reducing to 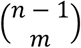 if *m* is known.

The 2nd to 4th order polynomial curve fitting are carried out to find best fit of data, to understand the curve around the daily confirmed cases along with forward projection. These polynomial fitting orders are computed based on following equations^35^;

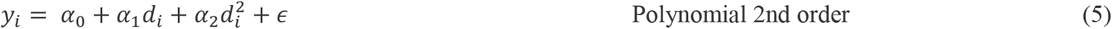

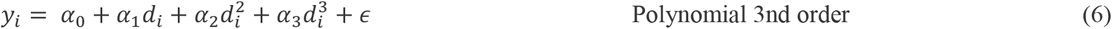

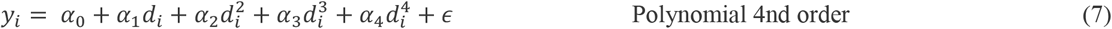

Where *y*_*i*_ is fitting curve function, *α*_0_ represent intercept, *α*_1_ to *α*_4_ are slope coefficients and *d*_*i*_ is function of *x* called basis function which is defined as;

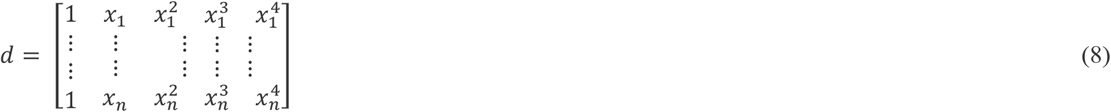

### The CoVID-19 status

The CoVID-19 pandemic displayed dichotomous global CIR ranging from 0 - 3.63% (28th January to 22nd February, 2020) that upsurge to 9.89% on 23rd March, 2020 (Fig. 1). Western pacific region represent dominantly towards global CIR up to 100% till 20th February, 2020. European region appeared to be 2nd region as hotspot area by contributing 28% towards daily global CIR on 23rd February, 2020; with maximum CIR of 84.04% on 13th March 2020. Eastern Mediterranean region observed as third region effected by CoVID-19 with CIR of 33.88% on 3rd March 2020; maximum CIR 45.47% on 12th March, 2020. The American region appeared fourth as hot spot with CIR of 17.36% on 12th March, 2020; with maximum CIR of 24.36% on 23rd March, 2020. The South East Asia and African region remained least affected due to pandemic with CIR of below 5% till 24th March, 2020. Western Pacific CIR started decreasing sharply on 20th February and reached below 5% on 13th March, 2020. However, global CRR showed delay with extended curves appeared around first week of February and reached maximum 8.89% on 24th March, 2020; owing to continuous contribution by Western Pacific region. Global above average affected countries except Pakistan across six regions are evaluated for CIR and CRR (Fig. 2, Extended Fig. 1). The China represented on average 55.2% of global infected population followed by Italy (12.0%), South Korea (9.0%), Iran (7%), USA (4.8%), Spain (4.7%), Germany (4%), France (2.6%) and rest of countries below 2.0%. China has shown sharp CoVID-19 recovery rate with CIR value of 88% followed by Italy and Iran around 5% and rest of countries recorded below 2%; owing to delayed onset of local transmission. Pakistan showed CIR of below 0.1% due to emergence of first case on 27th February, 2020 and CRR closed to zero as recovery cases started from 8th March, 2020.

**Fig. 1:**
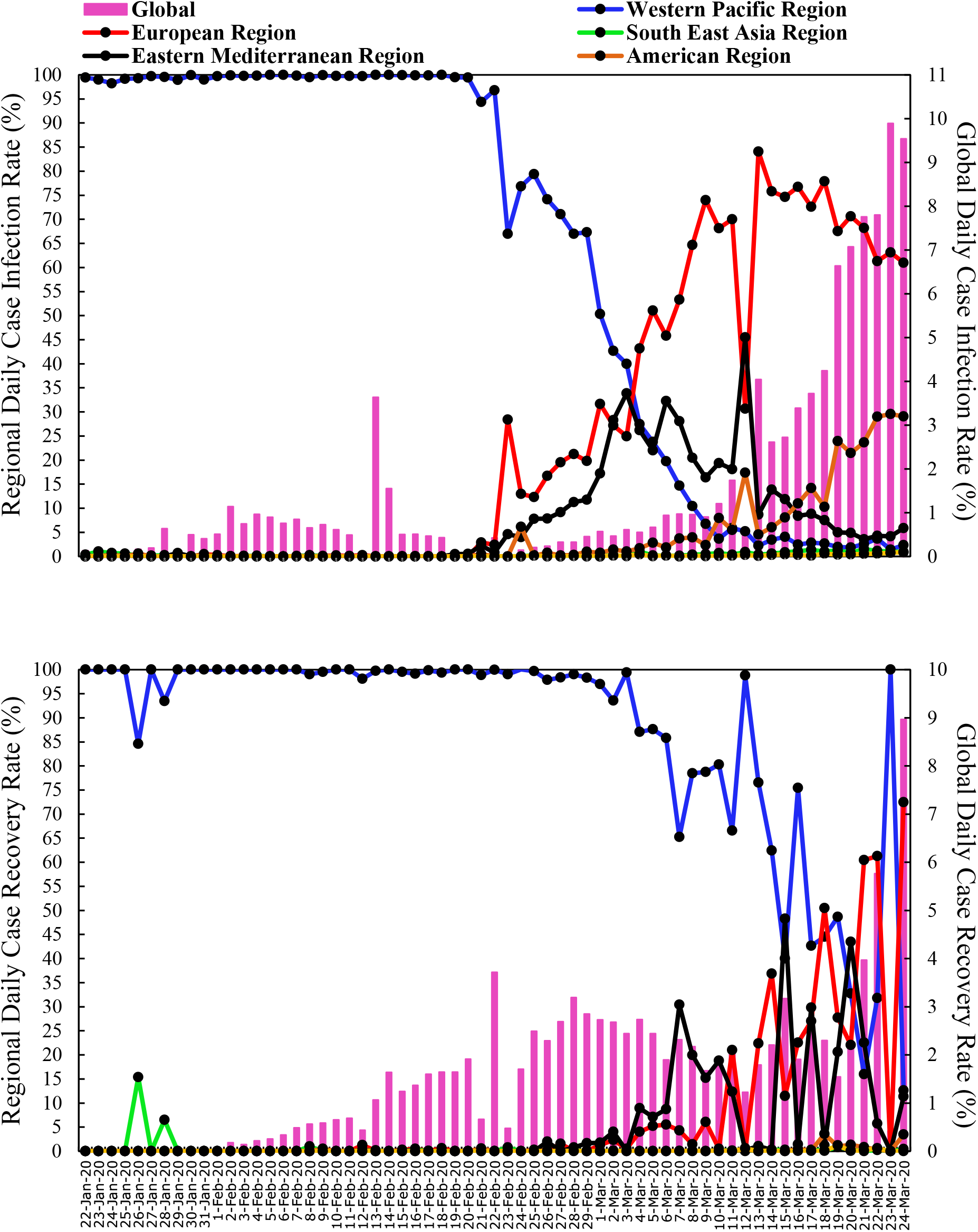
The CoVID-19 Global and Regional CIR and CRR highlighting associated temporal patterns.

**Fig. 2:**
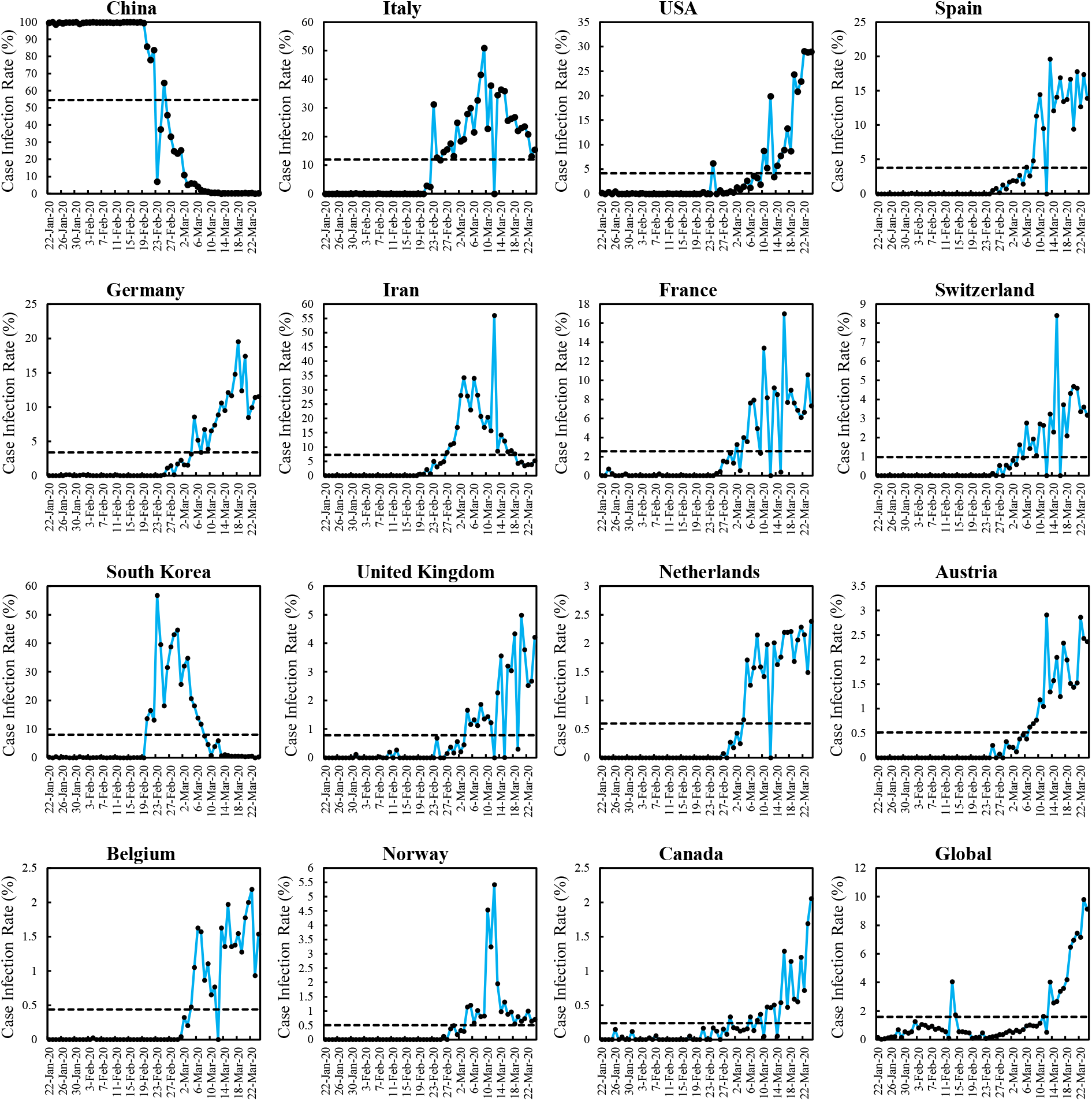
The CIR across above average selected countries along with mean (dash black line)

### Understanding CoVID-19 Pandemic Curve

To understand the pandemic curve, we evaluated time series data of confirmed cases based on statistical approach.

Probability density function along with measures of skewness and kurtosis displayed distinctive patterns for daily confirmed and recovered cases at regional level (Extended Fig. 2 & 3). All regions confirmed cases are high positive skewed (Skewness = 1.6 to 4.3) except for bimodal approximate symmetrical Eastern Mediterranean region (skewness = 0.4). Leptokurtic high and sharp peaks are observed for Western Pacific region (kurtosis = 26.6; mean = 1541), followed by American (kurtosis = 6.7; mean = 1011), South East Asia (kurtosis = 5.2; mean = 50), African (kurtosis = 4.9; mean = 25) and European (kurtosis = 4.3; mean = 3539). Platykurtic lower with broader peak is only observed for Eastern Mediterranean region (kurtosis = 6.7; mean = 454). The global confirmed cases showed highly positive skewed (skewness = 2.2) and leptokurtic (kurtosis = 6.8; mean = 6620) peak with extreme values on the right side tail.

Regions recovered cases are highly positively skewed (Skewness = 1.5 to 3.7) except for bimodal moderately skewed Western Pacific region (skewness = 0.6). Leptokurtic high and sharp peaks are observed for European region (kurtosis = 17.1; mean = 315), followed by South East Asian (kurtosis = 15.0; mean = 2), American (kurtosis = 14.6; mean = 9), African (kurtosis = 4.9; mean = 1) and Eastern Mediterranean (kurtosis = 4.6; mean = 150). Approximate Mesokurtic peak is only observed for Western Pacific region (kurtosis = 2.7; mean = 1231). The global recovered cases showed high positive bimodal skewed (skewness = 2.0) and leptokurtic (kurtosis = 10.0; mean = 1708) peak with less extreme values on the right side tail.

The pandemic curve is further evaluated to detect the multiple change point in time series to identify the phases and associated length period at selected countries, regions and global scale (Fig. 4, Extended Fig. 4 to 8). Each curve showed three phases of change along with last curve segment length. The first phase represent the hidden CoVID-19 incubation time period which indirectly represent emptiness of policy based actions to surround the spread. The second phase represent the exponential rise time and conversion into high risk for disease spread. Third phase signify disastrous impact on community health and economy. The last segment length denote time taken for the flattening of curve through lockdown, social distancing and other policy actions to curtail the exponential growth of pandemic event. Longer and lower last segment represent decline of the CoVID-19 curve.

**Fig. 3:**
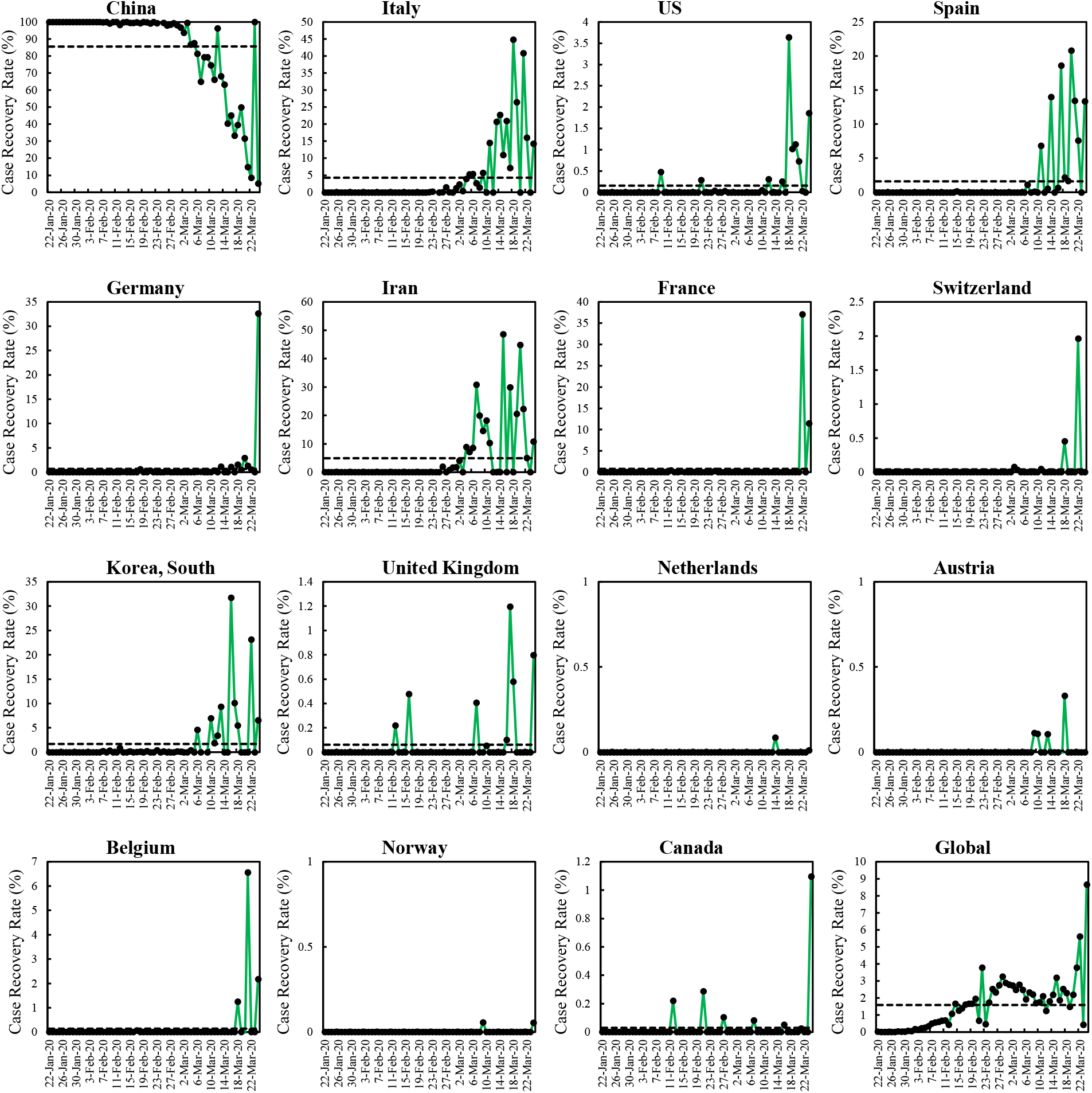
The CRR across above average selected countries along with mean (dash black line)

**Fig 4:**
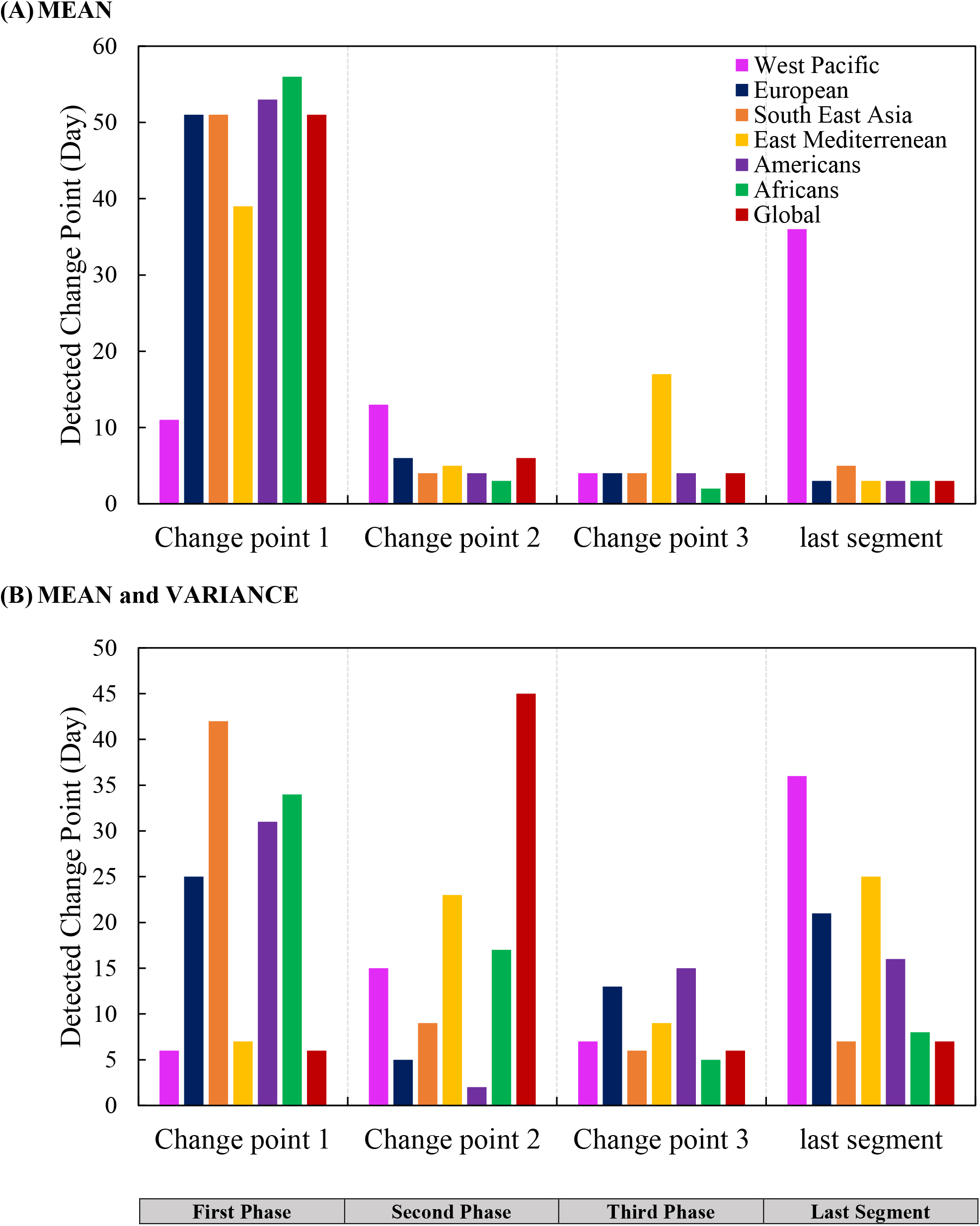
Multiple CPD based CoVID-19 three phases and last segment at regional scale during 64 days of global pandemic

The mean based CPD revealed first phase length remained 39 to 56 days (22nd January to 17th March, 2020) for all regions except for Western Pacific with 12 days (22 January to 1 February, 2020). The sharp rise in cases are observed for African (3 days), American and South East Asia (4 days), East Mediterranean (5 days), European (6 days) and Western Pacific (13 days) during second phase. The third phase remained in range of 2 to 4 days for regions except for Eastern Mediterranean region with 17 days. Length of last segment remained in range of 3 to 5 days except for Western Pacific region with 36 days representing pandemic cool down period. The global pandemic curve characteristics include 51 days, 6 days, 4 days and 3 days from first phase to last segment of the curve. The mean CPD ignore temporal variance of the data. So, the mean and variance CPD provide in-depth information above CoVID-19. The first phase length was found shorter for Western Pacific and East Mediterranean region with 6 to 7 days (22nd January to 28th January, 2020) followed by European (25 days), American (31 days), African (34 days) and South East Asia (42 days on 3rd March, 2020). All regions varied in term of second phase period and sharp rise in cases are observed for American (2 days) followed by European (5 days), South East Asia (9 days), Western Pacific (15 days), African (17 Days) and Eastern Mediterranean (23 days). The third phase displayed that African region is facing least impact due to pandemic (5 days) followed by South East Asia (6 days), Western Pacific (7 days), Eastern Mediterranean (9 days), European (13 days) and being highest for American (15 days). Length of last segment period remained in range of 7 to 25 days except for Western Pacific region with 36 days. The global pandemic curve characteristics include 6 days, 45 days, 6 days and 7 days from first phase to last segment of the curve. Further details for CPD related to affected countries are available in supplementary information.

The CPD help to understand the CoVID-19 curve in term of sudden change but lacks information about temporal trends and provide short term projections related to infected population. We attempted to evaluate simple and robust approach of polynomial curve fitting to estimate and project infected population (Fig. 5, Extended Fig. 9 to 10). Global infected population of 0.373 million confirmed cases is estimated with 2rd order polynomial (R^2^ = 0.87) that will increase to 0.750 million (R^2^ = 0.87), 1.900 million (R^2^ = 0.96) and 4.680 (R^2^ = 0.99) with 2nd, 3rd and 4th order polynomials, respectively by 24 April, 2014. The projected infected population remained in the range of 0 to 0.700 million (Western Pacific), 0.500 to 2.650 million (European), 0.006 to 0.048 million (South East Asia), 0.080 to 0.150 million (Eastern Mediterranean), 0.120 to 0.990 million (American) and 0.003 to 0.0230 million (African). The selected countries projections remained in the range of 0.000 to 0.625 million (China), 0.180 to 0.690 million (Italy),

**Fig. 5:**
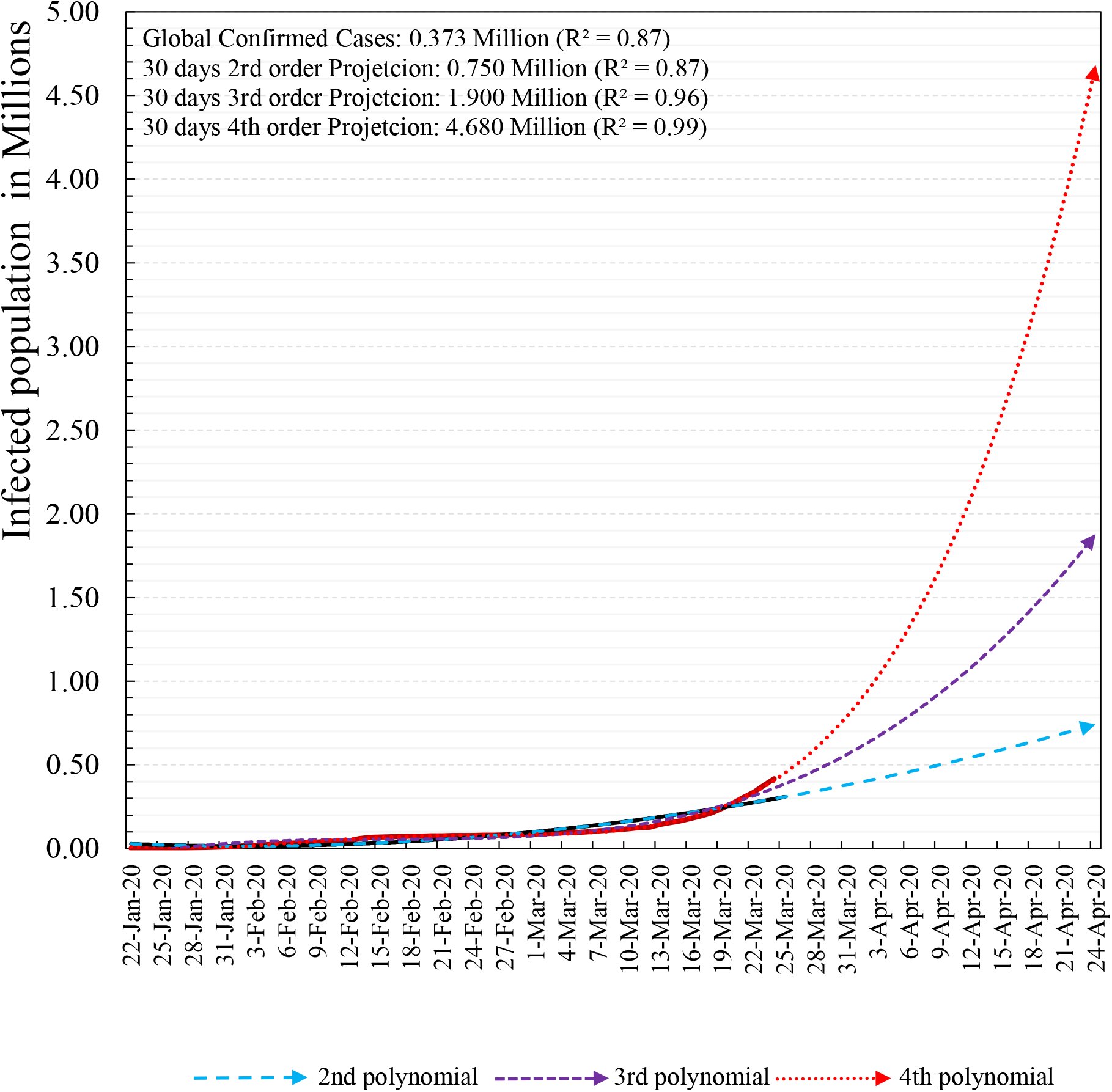
Global projected infected population based on 2nd, 3rd and 4th order polynomial curve fitting.

0.115 to 0.960 million (USA), 0.095 to 0.600 million (Spain), 0.080 to 0.480 million (Germany), 0.072 to 0.128 (Iran), 0.050 to 0.300 (France), 0.024 to 0.145 (Switzerland), 0.000 to 0.025 (South Korea), 0.019 to 0.125 million (UK), 0.012 to 0.076 million (Netherlands), 0.012 to 0.076 million (Austria), 0.010 to 0.061 million (Belgium), 0.008 to 0.025 (Norway), 0.006 to 0.044 million (Canada) and 0.002 to 0.016 (Pakistan).

The polynomial approach is assessed to determine best fitting order for CoVID-19 estimation and projection based on percent error (Table 1). The WHO situation report confirmed cases of 1st April, 2020 is used to evaluate the infected population projection for all three polynomial order. The 2nd order polynomial is found best in predicting infected population for Western Pacific region (R^2^ = 0.97) and China (R^2^ = 0.96) with error ranging between −15.7 to −10.7%. The infected population is best projected for Eastern Mediterranean region (R^2^ = 0.99), Italy (R^2^ = 0.88), Germany (R^2^ = 0.94), Iran (R^2^ = 0.99), Switzerland (R^2^ = 0.94), South Korea (R^2^ = 0.95), and Pakistan (R^2^ = 0.90) with error between −11.6 to 13.4% with 3^rd^ polynomial fitting. Rest of regions and countries is best projected by 4^th^ order polynomial with error of −26.3 to11.2% i.e., European region (R^2^ = 0.99), South East Asia (R^2^ = 0.98), American (R^2^ = 0.94) and African (R^2^ = 0.95), USA (R^2^ = 0.95), Spain (R^2^ = 0.99), France (R^2^ = 0.99), UK (R^2^ = 0.98), Netherlands (R^2^ = 0.99), Austria (R^2^ = 0.99), Belgium (R^2^ = 0.99), Canada (R^2^ = 0.96) and Pakistan (R^2^ = 0.97).

**Table 1:** Assessment of projected infected population on 1st April, 2020 at global, regional and selected countries scale.

| Scale | Confirmed<br>Cases (Million) | Projected Infected<br>population (Million) |  |  | Percent Error |  |  |
| --- | --- | --- | --- | --- | --- | --- | --- |
|  | 1st April 2020 | 2nd<br>order | 3rd<br>order | 4th<br>order | 2nd<br>order | 3rd<br>order | 4th<br>order |
| Global | 0.824 | 0.400 | 0.600 | 0.900 | -51.4 | -27.2 | **9.3** |
| WPR | 0.106 | 0.095 | 0.090 | 0.140 | **-10.7** | -15.4 | 31.6 |
| ER | 0.464 | 0.200 | 0.350 | 0.500 | -56.9 | -24.6 | **7.7** |
| SEAR | 0.007 | 0.003 | 0.006 | 0.008 | -58.4 | -16.8 | **10.9** |
| EMR | 0.054 | 0.036 | 0.048 | 0.047 | -33.7 | **-11.6** | -13.4 |
| AMR | 0.189 | 0.040 | 0.080 | 0.180 | **-78.8** | -57.6 | **-4.6** |
| AFR | 0.004 | 0.001 | 0.002 | 0.003 | -75.4 | -50.9 | **-26.3** |
| China | 0.083 | 0.070 | 0.065 | 0.120 | **-15.3** | -21.3 | 45.2 |
| Italy | 0.106 | 0.080 | 0.120 | 0.140 | -24.4 | **13.4** | 32.3 |
| USA | 0.163 | 0.040 | 0.080 | 0.150 | -75.5 | -51.0 | **-8.1** |
| Spain | 0.094 | 0.040 | 0.075 | 0.100 | -57.6 | -20.6 | **5.9** |
| Germany | 0.067 | 0.040 | 0.060 | 0.080 | -40.6 | **-10.9** | 18.8 |
| Iran | 0.045 | 0.026 | 0.044 | 0.040 | -41.7 | **-1.4** | -10.3 |
| France | 0.051 | 0.020 | 0.040 | 0.055 | -61.1 | -22.3 | **6.8** |
| Switzerland | 0.016 | 0.009 | 0.016 | 0.024 | -44.1 | **-0.7** | 49.0 |
| South Korea | 0.010 | 0.014 | 0.010 | 0.002 | 41.6 | **1.1** | -79.8 |
| UK | 0.025 | 0.008 | 0.014 | 0.021 | -68.2 | -44.3 | **-16.5** |
| Netherlands | 0.013 | 0.006 | 0.010 | 0.014 | -52.4 | -20.6 | **11.2** |
| Austria | 0.010 | 0.004 | 0.008 | 0.011 | -60.7 | -21.4 | **8.0** |
| Belgium | 0.013 | 0.004 | 0.008 | 0.011 | -68.7 | -37.4 | **-13.9** |
| Canada | 0.008 | 0.002 | 0.004 | 0.007 | -74.0 | -48.0 | **-9.0** |
| Pakistan | 0.002 | 0.001 | 0.002 | 0.002 | -51.0 | **-1.9** | **-1.9** |

The polynomial fitting also support in understanding the curve status and its association with socio-economic policy decisions. The 2^nd^ order polynomial association with Western Pacific region especially for China explained that CoVID-19 pandemic wave 1 has passed its peak and subsided to its lowest level. While, all regions and countries best fitted with 3rd order polynomial have reached closer to its peak; due to delayed policy action or lateral spread through local transmission. The 4th order polynomial refers where disease spread is at its initial stage and expected to grow exponentially; requiring strict policy action to curtail epidemic.

## Conclusions

Main findings are as follows:

- The CoVID-19 pandemic showed similar level of daily cases infection and recover rate up to 9.0 percent at global level.
- To flatten the pandemic curve, probability density function along with skewness and kurtosis measures provided insight to curve shape and height of its peak.
- The change point detection based on observed infected population helped to interpret disease curve into three distinctive phases. The China is the first country, where first wave of pandemic has reached to its end. Rest of countries are still active areas and time of its peak, infected population size will depend on effectiveness of community supported national action policies such that social distancing, health system capacity, access to food and medicine and others.
- Polynomial approach is used due to its ability to explain the non linear CoVID-19 curve. The 2nd order polynomial is found good for saturated disease curve like China. The pandemic curve at initial to peak stage are best estimated using 3rd and 4th order polynomials due to exponential growing infected population and lateral spread nature of the disease.
- Polynomial based infected population projection are subject to high risk due to daily changing situation and repeated projection at 3-7 days are necessary to control the curve.

## Data Availability

Data and codes are available on reasonable request.

## Acknowledgement

This research acknowledges the World Health Organization who made available the good quality data for this research.

